# Antigenic seniority and convergent haemagglutinin evolution shaped the immune landscape preceding influenza B/Yamagata’s extinction

**DOI:** 10.64898/2026.05.17.26353389

**Authors:** R. Steventon, M. Lee, R. Gregory, L. Stolle, C. McInally, L. Jarvis, N.C. Robb, G. Carnell, N. Temperton, S. Cherny, U. Obolski, C P. Thompson

**Author notes:** Corresponding authors &.

## Abstract

**Background:** In 2020, the B/Yamagata lineage of influenza B disappeared from circulation. Understanding the immunological conditions preceding this loss can help explain why the lineage disappeared and has implications for our understanding of viral evolution and ongoing vaccine policies.

**Methods:** We measured neutralising antibody responses in age- and sex-matched blood donor cohorts collected in 2020, 2023, and 2025 (n=114 per cohort) against a panel of pseudotyped influenza B viruses spanning 79 years of evolution, validated against live virus neutralisation assays. We performed antibody pulldown assays using full-length and head domain HA proteins from B/Yamagata (B/Phuket/3073/2013) and B/Victoria (B/Washington/02/2019) lineages, testing purified antibodies against chronologically distinct pseudotyped viruses. We used Bayesian network analysis, and LASSO regression to identify potential molecular determinants of cross-lineage neutralisation which were then confirmed via site-directed mutagenesis, and epitope-specific peptide pulldowns.

**Findings:** In 2020, population immunity was asymmetrically focused on B/Yamagata viruses. Despite no B/Yamagata circulation after 2020, neutralisation of recent B/Yamagata strains increased in 2023 while B/Victoria responses remained unchanged, consistent with antigenic seniority directing recall responses toward B/Yamagata. This trend inverted by 2025, with B/Yamagata immunity declining and B/Victoria immunity increasing. Antigen-specific purified antibodies showed lineage-biased potency, neutralising B/Yamagata viruses more effectively than B/Victoria viruses. We identified the 120-loop of the HA head domain as a critical cross-lineage epitope, with the charge state at a single convergently evolving residue at position 131 determining cross-reactive potency between lineages.

**Interpretation:** B/Yamagata’s acquisition of a positively charged residue at position 131, shared with B/Victoria, likely increased its cross-neutralisation by Victoria-raised immunity preceding its disappearance. Although antigenic seniority sustained cross-reactive responses to B/Yamagata after its extinction, the waning of this effect by 2025 suggests that population immunity to B/Yamagata is now declining. This has implications for influenza B vaccine policy.

**Funding:** CPT and UO were funded by a joint grant by the British Council and the Israeli Ministry of Innovation, Science & Technology, under the ISPF scheme [grant number 47650215]. N.C.R. is supported by a Royal Society Dorothy Hodgkin Research Fellowship [grant number DHR00620]. RG was funded by The Institute for Global Pandemic Planning at the University of Warwick, UK, as part of a philanthropically supported doctoral programme. M.L. was funded via the Medical Research Council doctoral training programme grant.

**Evidence before this study:** We searched PubMed and bioRxiv for articles published between Jan 1, 2010 and Apr 1, 2026, using the terms ‘influenza B’, ‘B/Yamagata’ or ‘B/Victoria’ combined with ‘cross-reactivity’, ‘extinction’, ‘disappearance’ or ‘antigenic seniority’, without language restrictions. Evidence quality was assessed by study design, reproducibility across independent datasets, and sample size. Assessment of these manuscripts confirmed that prior work had established that cross-lineage neutralising antibodies exist between influenza B lineages. Vaccination studies in humans and mice demonstrated that these cross-reactive responses are asymmetrically focused on B/Yamagata. Studies of first-infection history suggested that the initial lineage encountered can shape lifelong antibody profiles through antigenic imprinting. Global surveillance confirmed B/Yamagata’s absence from circulation after 2020, prompting its removal from quadrivalent vaccines. However, whether this asymmetry observed in vaccination settings extends to population-level immunity shaped by natural infection, the specific molecular determinants of cross-lineage neutralisation, and how population immunity has evolved since B/Yamagata’s disappearance remained unknown.

**Added value of this study:** Using age-and sex-matched blood donor cohorts from 2020, 2023 and 2025, we demonstrate that population immunity was asymmetrically focused on B/Yamagata in 2020, widening further by 2023 consistent with antigenic seniority. An inversion of this pattern was seen by 2025 as B/Yamagata-specific responses waned. Among singleton positions unambiguously identifiable by LASSO, position 131 showed the strongest temporal increase, governing cross-reactive potency. B/Yamagata’s evolution to a positively charged residue at this position, matching the charge state of B/Victoria, likely increased its vulnerability to Victoria-raised immunity in the years preceding its disappearance.

**Implications of all the available evidence:** The waning of antigenic seniority-reinforced immunity to B/Yamagata by 2025 indicates that population protection against this lineage is declining. Should a B/Yamagata-like virus re-emerge, the population may face substantially reduced lineage-specific protection. Ongoing serological surveillance will be needed to determine when this declining immunity reaches a threshold warranting reconsideration of vaccine composition.

## Introduction

In early 2020, the B/Yamagata lineage of influenza B virus disappeared from global circulation^1,2^. This lineage had co-circulated with B/Victoria for two decades, each accounting for approximately half of seasonal influenza B cases worldwide^3,4^. B/Yamagata has not been detected since the COVID-19 pandemic began, representing an unprecedented disappearance of a major human respiratory pathogen^5^. Subsequently, from September 2023, the WHO recommended that the B/Yamagata strain be removed from the seasonal influenza vaccine for both the Northern and Southern Hemisphere^6^. Consequently, understanding why B/Yamagata vanished, and the impact that this loss has on population immunity to influenza B, has critical implications for virology and vaccine policy

The two influenza B lineages emerged sequentially: B/Victoria in approximately 1972, and B/Yamagata in the late 1980s^7^. They established stable global co-circulation after 2000^4,8^. Unlike influenza A, which maintains multiple subtypes through animal reservoirs, influenza B circulates exclusively in humans^9,10^ making lineage eradication theoretically possible through population immunity alone. Non-pharmaceutical interventions due to the COVID-19 pandemic, intrinsic viral factors such as antigenic conservation, and depleted susceptibility following an outbreak in 2017/2018 have been proposed to explain B/Yamagata’s disappearance in 2020^11–13^.

Several studies provide evidence supporting the hypothesis that asymmetric cross-lineage immunity may have also contributed to B/Yamagata’s disappearance^14^. Despite antigenic distinction between lineages, cross-lineage antibodies between B/Yamagata and B/Victoria have been shown to exist with unusual properties. A subset of haemagglutinin (HA) targeting, cross-lineage antibodies have previously been shown to preferentially neutralise B/Yamagata over B/Victoria viruses^15^, and pre-existing B/Yamagata immunity has been shown to block subsequent B/Victoria vaccine responses, while the inverse does not occur^16,17^. Recent animal studies have also implicated neuraminidase-specific antibodies in asymmetric cross-lineage protection^18^. This asymmetry may reflect distinct evolutionary constraints with B/Yamagata maintaining multiple co-circulating clades, whilst B/Victoria evolves along a single continuous backbone exhibiting greater antigenic evolution^19,20^. Together, this evidence supports the theory that increased population immunity to conserved epitopes, which enable cross-lineage recognition, may have antigenically constrained B/Yamagata, rendering it increasingly vulnerable to extinction as cross-protective immunity accumulated over time, although population-level evidence has yet to substantiate this mechanism.

Consequently, critical gaps remain regarding our understanding of the extent and distribution of cross-lineage neutralising antibodies within human populations, the role of conserved versus variable epitopes in mediating cross-protection, and whether pre-existing immunity patterns created sufficient selective pressure to eliminate B/Yamagata in 2020. Here, we address these questions through comprehensive serological analysis of age-stratified human sera against historical and contemporary viruses from both influenza B lineages. We demonstrate that asymmetric population immunity and antigenic seniority were associated with the immunological conditions in 2020 which preceded B/Yamagata’s disappearance. Surprisingly, we show that despite B/Yamagata not circulating post-2020, antibody-mediated population immunity to it still increased in 2023, whilst immunity to B/Victoria lineages viruses remained constant. In 2025, this trend inverted with B/Yamagata levels dropping and B/Victoria levels increasing. Furthermore, using single-infection ferret antisera and antibody pulldowns with full-length and head domain HA antigens, we demonstrate that cross-lineage antibodies preferentially neutralise historic B/Yamagata viruses over historic B/Victoria viruses. Finally, we identify a region of the 120-loop of the HA head domain as a critical epitope contributing to the observed inter-lineage cross-reactivity.

Collectively, our findings define an immunological mechanism that may have contributed to the disappearance of B/Yamagata, provide evidence that antigenic seniority has shaped the antigenic landscape of influenza B, and highlight how population immune structure influences viral survival, competition and extinction. We propose that the asymmetric population immunity directed toward B/Yamagata reflects the distinct evolutionary trajectories of the two lineages: B/Yamagata historically maintained multiple co-circulating clades, enabling successive infections to repeatedly boost pre-existing memory B cells and reinforce the antigenic seniority of shared epitopes, whereas B/Victoria evolved along a single continuous backbone, necessitating recruitment of naïve B cells to target newly emerged epitopes. Furthermore, we identify a convergent antigenic evolution at a critical epitope within the 120-loop whereby B/Yamagata’s acquisition of a positively charged residue, shared with B/Victoria, rendered it increasingly susceptible to cross-neutralisation by Victoria-raised immunity. The persistence of antigenic seniority–reinforced antibody responses to B/Yamagata following its disappearance, together with the recent waning of this effect, has important implications for vaccine policy.

### Population immunity was asymmetrically focused on B/Yamagata in 2020 and shifted following its disappearance

To examine the level of asymmetric immunity towards B/Yamagata viruses within the population, neutralising antibody responses were measured in cohorts of 114 blood donor sera obtained in 2020, 2023, and 2025, spanning the period of B/Yamagata’s circulation and subsequent disappearance. The cohorts were matched for age and sex (Supplementary Figure 1). These sera were run against a panel of 9 pseudotyped viruses representing 79 years of viral evolution (Supplementary Table 1). This panel included one ancestral strain, four B/Victoria-like strains, and four B/Yamagata-like strains. A neutralisation threshold was established as five standard deviations above the mean response using an Ebola glycoprotein pseudotyped virus control (Supplementary Figure 2a). Pseudotype microneutralisation (pMN) results were validated by live virus neutralisation assays, demonstrating strong correlations between pseudotype and live virus for both a B/Yamagata and a B/Victoria influenza B virus (Supplementary Figure 2b).

In 2020, neutralisation titres against B/Yamagata-like viruses were significantly higher than all B/Victoria-like viruses tested except for B/Florida/4/2006 (Figure 1a and Supplementary Figure 3a). To ensure that the output from our pMN assays was due to the HA sequence and not the abundance of HA differing between viruses, a western blot was run to show that the pseudotyped viruses displayed equivalent amounts of HA on their surfaces (Supplementary Figure 2c). This asymmetric immune profile towards the B/Yamagata lineage widened in 2023 despite B/Yamagata not circulating since 2020 with neutralisation of B/Yamagata/16/1988 and B/Phuket/3073/2013 increasing, although no statistically significant increase was seen for B/Hong Kong/03/1992 or B/Florida/4/2006. Surprisingly, no statistically significant increase was detected for any of the B/Victoria pseudotyped viruses: B/Victoria/2/1987, B/Bangkok/163/1990, B/Hong Kong/330/2001 and B/Washington/02/2019 (Figure 1a).

**Figure 1.**
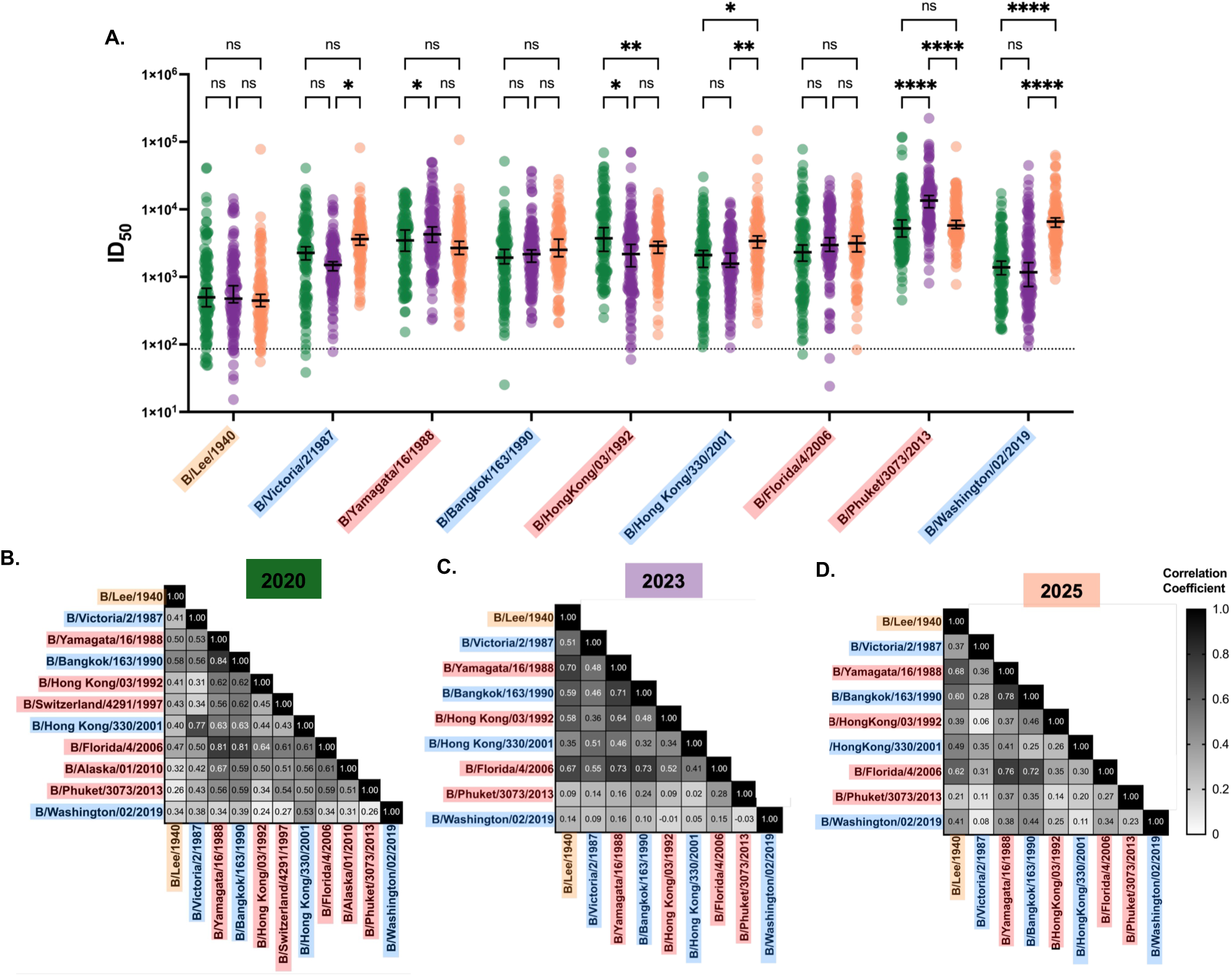
Pseudotype microneutralisation (pMN) of influenza B viruses across three donor cohorts reveals asymmetric immunity. Sera (n = 114) from Scottish blood donors collected in 2020 (green), 2023 (purple), and 2025 (orange) were tested against a panel of influenza B pseudotype viruses. Victoria-lineage viruses are shown in blue, Yamagata-lineage viruses in red, and the ancestral B/Lee/1940 virus in orange. **(A)** Neutralising antibody titres (ID_50_) against historical and contemporary influenza B viruses. The dotted line indicates the lower limit of detection. Bars represent median values with 95% confidence intervals. Statistical comparisons were performed using two-tailed Mann–Whitney U tests with Dunn’s correction for multiple comparisons (*p < 0.05, **p < 0.01, ***p < 0.001, ****p < 0.0001). **(B–D)** Spearman’s rank correlation matrices of pairwise ID_50_ values for the 2020, 2023, and 2025 cohorts, respectively.

This trend inverted in 2025. Neutralisation of B/Victoria viruses increased for B/Victoria/2/1987, B/Hong Kong/330/2001 and B/Washington/02/2019 in 2025 versus 2023. Neutralisation of B/Hong Kong/330/2001 and B/Washington/02/2019 also increased in 2025 versus 2020. In contrast in 2025, neutralisation of B/Yamagata viruses decreased for B/Phuket/3073/2013 2025 vs 2023, and B/Hong Kong/03/1992 2025 versus 2020 or remained stable (Figure 1a). This resulted in a reduction in the asymmetric immune profile with neutralisation titres towards historic Victoria-like viruses no longer being significantly different to historic Yamagata-like viruses (Supplementary Figure 3b-c).

For all cohorts, correlation analysis showed that there were strong associations between B/Yamagata and B/Victoria viruses (classed as a Spearman’s rank correlations of >0.7). For example, B/Yamagata viruses, B/Florida/4/2006 and B/Yamagata/16/1988 strongly correlated with B/Victoria virus, B/Bangkok/163/1990 in all cohorts (Figure 1b-d).

Collectively, these data indicate that in March 2020, there was greater antibody mediated population immunity to B/Yamagata strains than B/Victoria strains. Even after the disappearance of the B/Yamagata lineage in 2020, antibody responses still increased against B/Yamagata but not B/Victoria lineage viruses, in a fashion consistent with antigenic seniority. This pattern persisted until 2025, when antibody responses to B/Victoria strains began to rise, while those to B/Yamagata declined or remained stable.

### Cross-lineage antibodies target HA head and stem domains with asymmetric neutralisation potency

As our data proposed the existence of asymmetric population immunity, we next investigated whether this was due to antibodies capable of directly cross-neutralising between lineages, and, if so, which HA domains they target. To address this, we assessed whether antibodies raised against contemporary B/Victoria viruses could neutralise historical B/Yamagata strains using ferret antisera. Antisera generated against three recent B/Victoria viruses, B/Slovenia/1584/2020, B/Stockholm/3/2022, and B/Netherlands/11267/2022, were tested for neutralisation against a panel of B/Victoria and B/Yamagata lineage pseudotyped viruses.

Neutralisation responses were highest against B/Washington/02/2019, the most antigenically similar virus in the panel, across all three ferret sera (Figure 2a). Notably, all antisera neutralised historical B/Yamagata viruses more effectively than historical B/Victoria strains. For example, antisera raised against B/Stockholm/3/2022 failed to yield measurable ID₅₀ values against older B/Victoria strains (B/Victoria/2/1987 and B/Hong Kong/330/2001) but readily neutralised B/Yamagata strains (B/Yamagata/16/1988 and B/Bangkok/163/1990). (Figure 2a).

**Figure 2.**
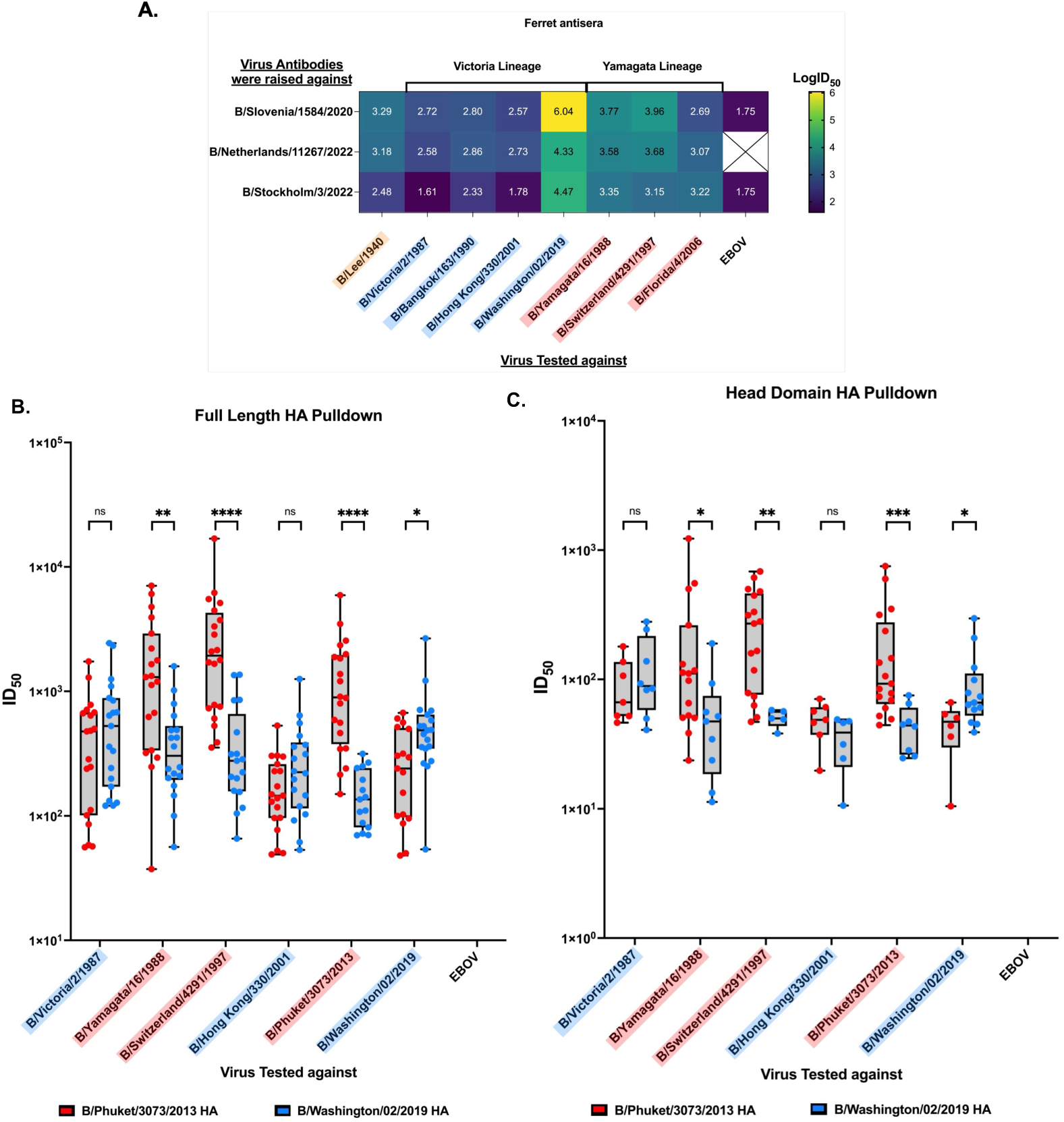
Cross-lineage neutralisation of influenza B pseudotype viruses. **(A)** Ferret antisera raised against three Victoria-lineage viruses were tested by pMN against Victoria-lineage, Yamagata-lineage, and ancestral pseudotype viruses. Titres are shown as log₁₀ ID_50_. The B/Netherlands11267/2022 was unresolvable. **(B–C)** Antibodies specific to the HA of B/Washington/02/2019 (Victoria) or B/Phuket/3073/2013 (Yamagata) were purified from 20 sera samples, then tested by pMN against a subset of Victoria-lineage and Yamagata-lineage pseudotype viruses. **(B)** Full-length HA used as pulldown antigen. **(C)** HA head domain used as pulldown antigen. Victoria-lineage viruses are indicated in blue and Yamagata-lineage viruses in red. Titres are expressed as ID_50_. Statistical comparisons were performed using two-tailed Mann–Whitney U tests with Dunn’s correction for multiple comparisons (*p < 0.05, **p < 0.01, ***p < 0.001, ****p < 0.0001).

To determine whether a similar asymmetry exists in humans, we performed antibody pulldown assays using full-length HA or isolated head domains from B/Victoria (B/Washington/02/2019) and B/Yamagata (B/Phuket/3073/2013) viruses as capture antigens^21^. Purified antibody fractions were then tested for neutralising activity using a pMN assay against a panel of historical and contemporary strains spanning both lineages. An EBOV pseudotyped virus control was included, as in Figure 2b&c.

Full-length HA pulldowns revealed that approximately 75% of sera contained cross-lineage neutralising antibodies (Figure 2b). However, their potency was asymmetric: antibodies captured with the B/Yamagata lineage HA antigen showed significantly higher ID₅₀ values against B/Yamagata lineage viruses than antibodies captured with the B/Victoria lineage antigen. In contrast, for historic B/Victoria lineage viruses, both antibody pulldowns performed equivalently. This asymmetry, in which overall B/Yamagata strains exhibit greater neutralisation, mirrors the population-level pattern whereby the antibody repertoire neutralises B/Yamagata lineage viruses more effectively.

Head domain pulldowns showed lower, but still detectable, cross-lineage activity in 25–45% of samples, indicating that cross-reactive epitopes are present within the immunodominant head as well as the more conserved stem (Figure 2c). The same asymmetric pattern persisted whereby antibodies captured via a B/Yamagata lineage HA antigen showed significantly higher ID₅₀ values against B/Yamagata lineage viruses than antibodies captured with the B/Victoria lineage antigen, but B/Victoria antigen pulldowns performed equally well against B/Yamagata and historic B/Victoria viruses.

Together, these findings demonstrate that cross-lineage antibodies target both HA stem and head domains, but with a consistent asymmetry in neutralisation potency that favours B/Yamagata lineage viruses, reflecting the broader population-level immune landscape shown in Figure 1.

### Position 131 within the 120-loop is a key determinant of cross-lineage antigenic variation

To identify the molecular basis of the cross-lineage serological relationships, we performed an Augmented Bayesian Network (ABN) analysis of the 2020 neutralisation data. As pseudotyped viruses display only influenza B HA, serological differences can be attributed to HA sequence variation.

The ABN revealed a serological network divergent from phylogeny (Supplementary Figure 4). B/Yamagata/16/1988 occupied the root position with cross-lineage dependencies, B/Washington/02/2019 was peripherally connected (weakest edge weight of β = 0.502), and B/Alaska/01/2010 was serologically isolated (Supplementary Figure 4). The cross-lineage connections, particularly B/Yamagata/16/1988 predicting responses to Victoria-like viruses, imply that shared HA sequence features drive these serological relationships.

To identify which HA residues contribute to these serological relationships, we employed LASSO (L1-penalised) regression with bootstrap stability selection, predicting pairwise serological similarity from residue similarity across variable HA1 positions (see Methods). Perfectly correlated positions - those with identical binary similarity patterns across all strain pairs - were grouped and represented by a single feature, yielding 34 independent position groups for the 11-strain analysis (Figure 3a, Supplementary Figure 5). Among the singletons - positions whose selection is unambiguous because their binary pattern is not shared with any other position - position 131 was selected in 13.0% of bootstrap resamples. Its four-state variation (N, H, K, and R) across the 11 strains makes it the most informationally rich singleton in the analysis (Figure 3a). Results were robust to alternative model specifications (Supplementary Table 2). Extension to the full-length HA showed minimal HA2 contribution, consistent with stem-directed antibodies mediating broad rather than strain-specific cross-reactivity (Supplementary Table 3).

**Figure 3.**
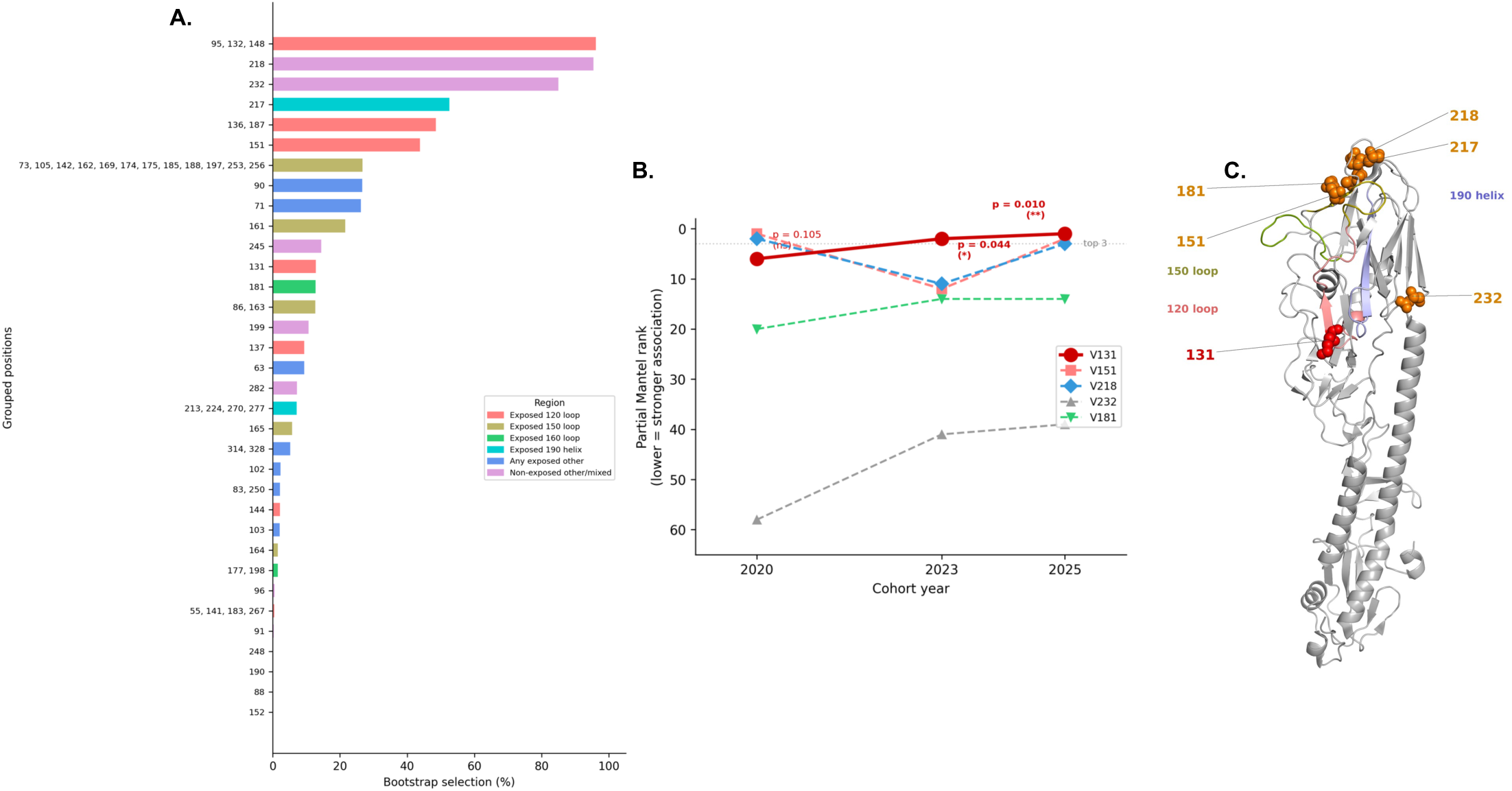
Grouped LASSO regression and partial Mantel tests identify position 131 as a key determinant of serological variation across influenza B viruses. **(A)** Grouped LASSO bootstrap stability for the 2020 cohort (n = 114 sera) against 11 influenza B pseudotyped viruses. Perfectly correlated positions — those with identical binary similarity patterns across all strain pairs — were grouped and represented by a single feature, yielding 34 independent position groups. Bars show the percentage of 1,000 bootstrap resamples in which each group was selected. Groups are colour-coded by antigenic region based on the first exposed position within the group: 120 loop (salmon), 150 loop (olive), 160 loop (green), 190 helix (cyan), any exposed other (blue), and non-exposed or mixed (pink). Antigenic site boundaries follow Ni et al. (2014): 120 loop (HA1 116–137), 150 loop (HA1 141–150), 160 loop (HA1 162–167), 190 helix (HA1 194–202). Position 131 is a singleton whose four-state variation (N, H, K, R) produces a unique binary pattern not shared with any other position. **(B)** Partial Mantel test ranking for singleton positions across three independent cohorts (2020, 2023, 2025; 9 common strains). Position 131 (red) shows a monotonic increase from rank 6 (p = 0.105) in 2020 to rank 1 (p = 0.010) in 2025, while positions 151 (pink) and 218 (blue) fluctuate without a consistent temporal trend. Lower rank indicates stronger partial association with serological similarity after controlling for all other variable positions. *p < 0.05, **p < 0.01. **(C)** Crystal structure of the B/Yamanashi/166/1998 HA monomer (PDB: 4M40) with antigenic regions coloured on the cartoon backbone: 120 loop (salmon), 150 loop (olive), 160 loop (green), 190 helix (blue). Singleton positions exceeding 50% bootstrap stability in at least one cohort year are shown as spheres: position 131 (red) within the 120 loop, and positions 151, 181, 217, 218, and 232 (orange)

To test reproducibility across independent cohorts, we repeated the grouped LASSO analysis on the 2023 and 2025 cohort data (n = 114 each) using the 9 strains common to all three timepoints (Supplementary Table 4). Position 131 showed a marked increase in bootstrap stability: 13.0% in 2020, 75.6% in 2023, and 74.1% in 2025 (Figure 3b). This temporal strengthening was independently confirmed by partial Mantel tests, in which position 131 rose from rank 6 (p = 0.105) in 2020 to rank 1 (p = 0.010) in 2025, while other singleton positions (151, 218) fluctuated without a consistent temporal trend (Figure 3c). Although position 151 also showed high bootstrap stability in 2025 (91.9%), its variation (E/K/R) is dominated by a single-strain outlier; all strains carry positively charged residues (K or R) except B/Washington/02/2019, which uniquely acquired glutamic acid (E). In contrast, position 131 shows genuine convergent evolution between lineages toward a shared positive charge, with B/Yamagata’s acquisition of lysine after 2015 aligning with its disappearance (Figure 4). The 95/132/148 group declined from 96.3% to 43.3% over the same period, consistent with a signal dominated by B/Washington/02/2019-specific variation that does not generalise as population immunity shifts.

**Figure 4.**
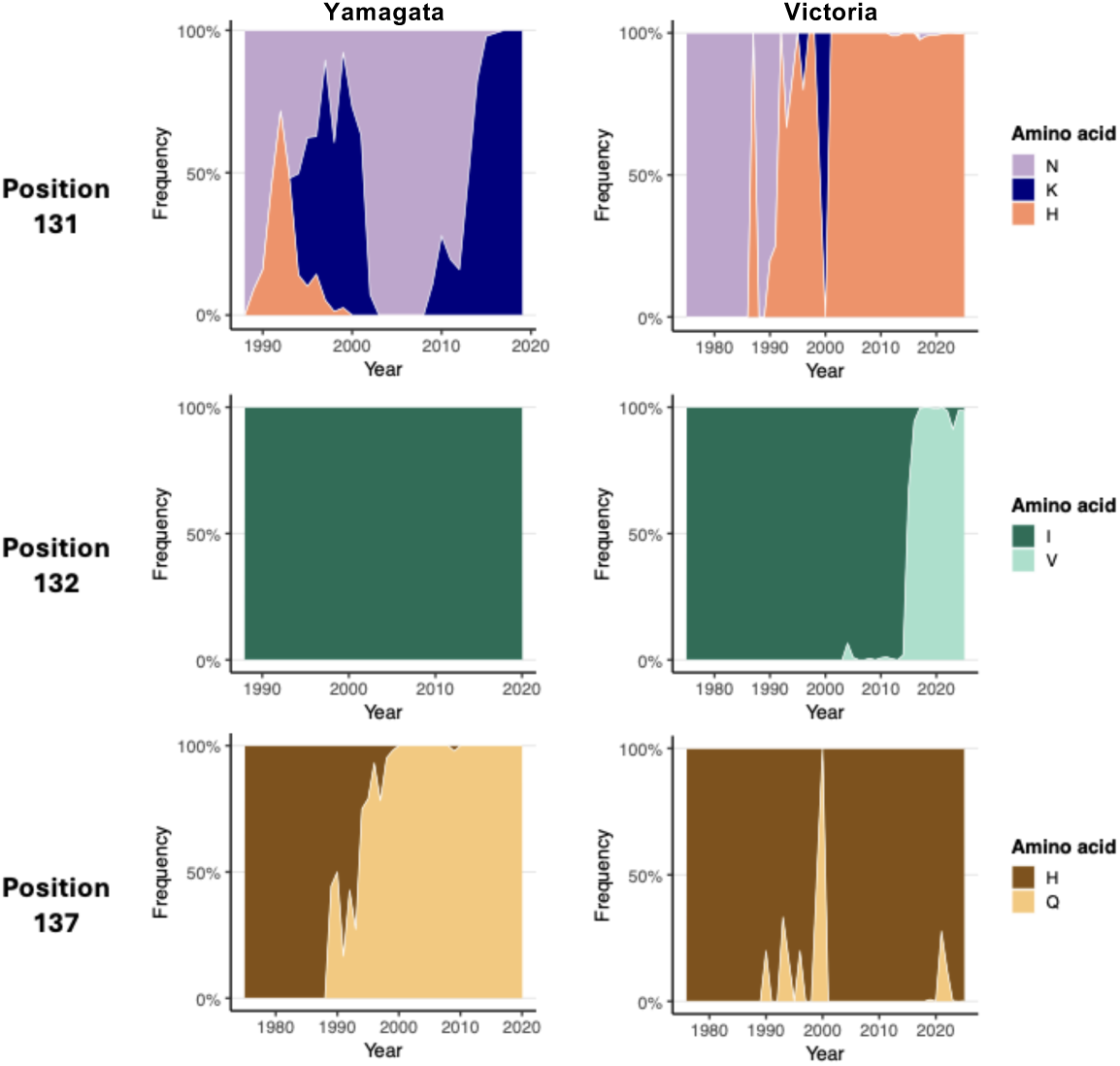
Evolutionary trajectory of amino acid variation at three co-varying positions within the 120-loop epitope. Frequency of amino acid variants at positions 131, 132, and 137 across influenza B HA sequences from GISAID (n = 80,000; 1940–2025), shown separately for B/Yamagata (left) and B/Victoria (right) lineages. At position 131, the ancestral asparagine (N, purple) was shared by both lineages until the early 1990s, when histidine (H, orange) emerged independently in both. B/Victoria maintained histidine from the late 1990s onwards, whereas B/Yamagata reverted to asparagine, which remained predominant until after 2015 when lysine (K, dark blue) became dominant. At position 132, both lineages retained the ancestral isoleucine (I, dark green) throughout most of their circulation history; B/Victoria acquired valine (V, light green) after 2014. At position 137, B/Victoria retained the ancestral histidine (H, brown), while B/Yamagata diverged to glutamine (Q, gold) by the late 1990s. Together, these trajectories produced the lineage-specific motifs at positions 131-132-137: H-V-H in B/Victoria and K-I-Q in late B/Yamagata.

Analysis of 80,000 GISAID sequences (1940–2025) revealed convergent evolution at the 120-loop (Figure 4). At position 131, the ancestral asparagine (N) was independently replaced by histidine (H) in both lineages during the early 1990s. B/Victoria maintained H thereafter, while B/Yamagata reverted to N before acquiring lysine (K) after 2015. All substitutions at position 131 (N to H, N to K, and the arginine present in B/Hong Kong/330/2001) represent convergent acquisition of positive charge. At position 137, B/Victoria retained the ancestral histidine while B/Yamagata diverged to glutamine (Q) by the late 1990s. Position 132 remained conserved as isoleucine until 2014, when B/Victoria acquired valine, producing a distinctive H-V-H motif at positions 131-132-137 in 82% of Victoria sequences. B/Yamagata retained the N-I-Q or K-I-Q motif (Figure 4).

### Natural inter-lineage variation at position 131 alters cross-reactive antibody recognition

To experimentally validate the computational identification of the 120-loop as a target of cross-reactive neutralising antibodies, we performed site-directed mutagenesis on a B/Switzerland/4291/1997 backbone carrying lysine at position 131. Alanine scanning across positions 130–138 (Figure 5a) revealed that K131A produced the largest reduction in neutralisation by human sera of any mutation tested (p < 0.0001), with individual sera showing up to 17-fold reductions in ID₅₀ (Figure 5b). Specific substitutions at position 131 confirmed its antigenic role: K131N significantly reduced neutralisation (p < 0.01), as did K131H (p < 0.05). Notably, K131H was the least disruptive substitution, consistent with the shared positive charge between lysine and histidine, whereas K131N, which replaces a positively charged residue with the uncharged ancestral asparagine, produced a larger reduction. When the uncharged asparagine was switched to the charged K and H within the B/Lee/1940 backbone, a significant reduction in neutralisation was also seen (Supplementary Figure 6). Adjacent positions E130 and R133 showed modest but significant effects upon alanine substitution. Q137A also significantly reduced neutralisation (p < 0.001), consistent with the co-varying role of position 137 identified by computational analysis. S135A, T136A, and N138A did not significantly alter neutralisation. Mutation of residues 132 and 134 failed to yield viable pseudotype viruses, potentially due to the structural importance of these residues. A control mutation outside the 120-loop (P270S) had no effect on neutralisation. Collectively, these results confirm that position 131 is a key antigenic determinant within the 120-loop and that the charge state at this position influences antibody recognition.

**Figure 5.**
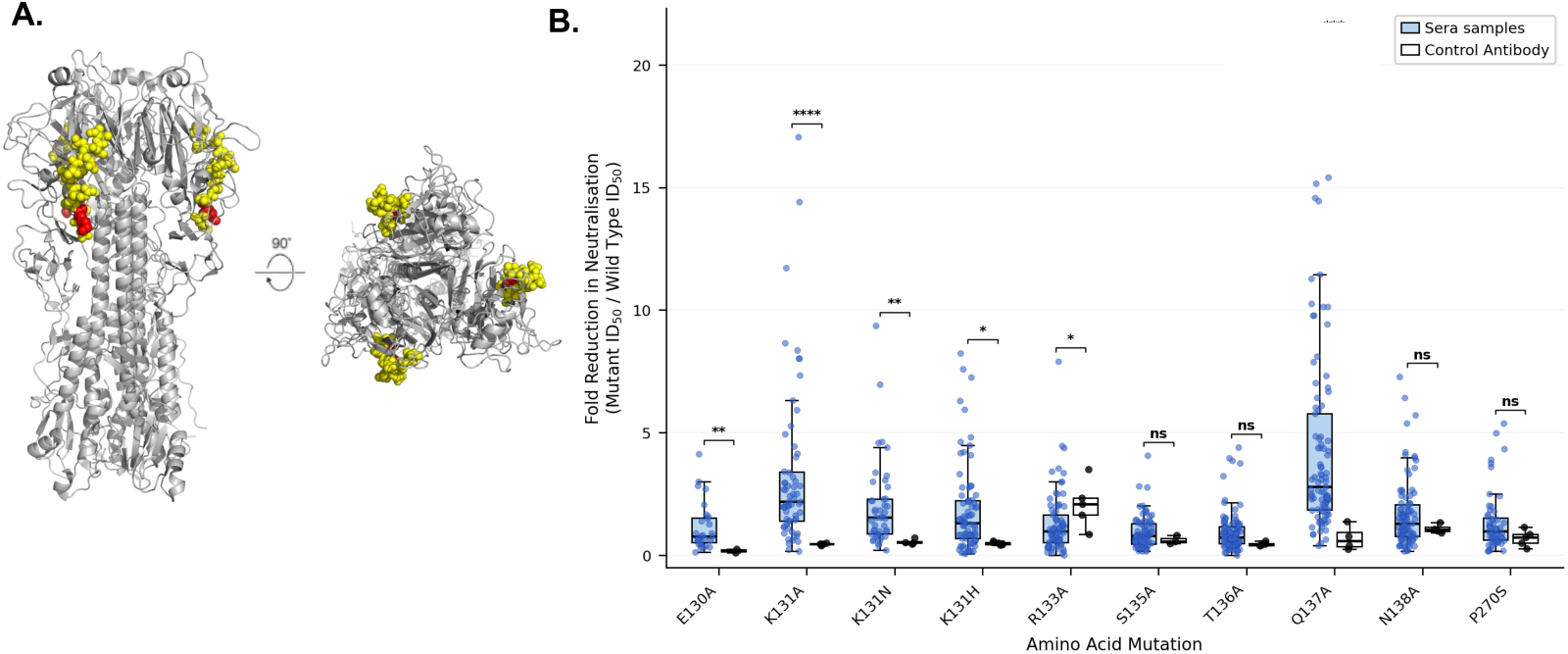
Site-directed mutagenesis validates the functional importance of 120-loop positions 131 and 137. **(A)** Crystal structure of B/Yamanashi/166/1998 HA showing the 120-loop (yellow) and position 131 (red). **(B)** Neutralisation of site-directed mutants by sera from the 2020 donor cohort (n = 114). Mutations were introduced into the B/Switzerland/4291/1997 HA background and tested by pMN assay. Data are shown as fold reduction in ID_50_ compared to wild-type virus. CR9114 monoclonal antibody was used as a control. Statistical comparisons were performed using two-tailed Mann–Whitney U tests with Dunn’s correction for multiple comparisons (*p < 0.05, **p < 0.01, ***p < 0.001, ****p < 0.0001).

### The 120-loop epitope mediates charge-dependent cross-lineage neutralisation

To characterise the antibody populations targeting the 120-loop epitope, we synthesised 20-mer peptides spanning positions 129–141 of HA1 representing the three major naturally occurring conformations at positions 131 and 137: HH (H at 131, H at 137; Victoria), NQ (N at 131, Q at 137; mid-Yamagata), and KQ (K at 131, Q at 137; late Yamagata)(Figure 6a). ELISA against individual sera (n = 19) confirmed that all sera contained antibodies recognising all three epitope conformations, with a significant hierarchy in binding (Friedman test, p < 0.0001): HH > NQ > KQ. This hierarchy is consistent with the circulation history of each conformation: B/Victoria has carried H at position 131 continuously since the late 1980s, providing the longest period of cumulative exposure, whereas the KQ conformation only became predominant in B/Yamagata after 2015 (Figure 6b-c).

**Figure 6.**
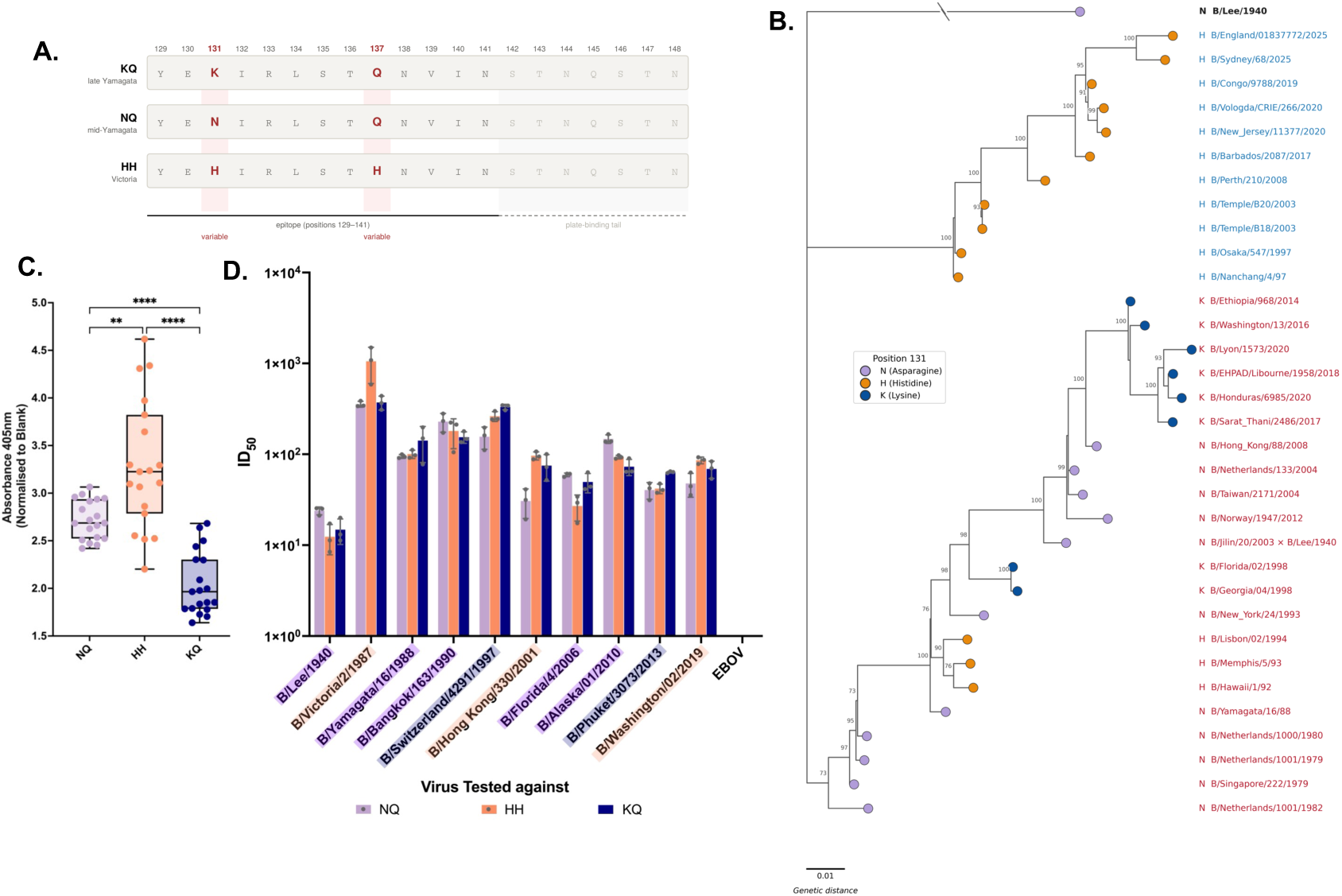
Position 131/137 variants within the 120-loop define functionally distinct epitopes with differential binding and neutralisation properties. **(A)** Sequences from the 120-loop epitope region showing amino acid variants at positions 131 and 137 used for peptide synthesis. **(B)** Maximum likelihood phylogenetic tree of influenza B viruses coloured by amino acid identity at position 131. Branch lengths represent genetic distance. Victoria-like viruses shown in blue, Yamagata-like viruses shown in red. **(C)** ELISA binding of donor sera (n = 20) to synthetic peptides representing the three major position 131 variants. Data shown as optical density at 405 nm. **(D)** Antibody pulldown assays using the same synthetic peptides. Antibodies specific to the position 131 variant peptides were pooled from 20 sera donors, then tested by pMN against pseudotype viruses bearing matching or mismatched 120-loop variants. Data represent biological triplicates and are shown as ID_50_ neutralisation titres.

We next purified antibodies from pooled human sera using the same peptides and tested their neutralising activity against a panel of pseudotyped viruses in triplicate (Figure 6d). Nine of ten viruses were most potently neutralised by antibodies purified with the peptide matching their residue at position 131, confirming that variation at this position defines epitope-specific antibody recognition. All three antibody populations demonstrated cross-lineage neutralisation; KQ-purified antibodies neutralised Victoria-like viruses as well as Yamagata-like viruses, and HH-purified antibodies neutralised Yamagata-like viruses as well as Victoria-like viruses, albeit at reduced potency compared to matched targets. The 120-loop epitope therefore mediates cross-lineage neutralisation, with position 131 determining the relative potency against matched versus mismatched viruses.

Examination of the cross-neutralisation patterns revealed that the charge state at position 131 was the principal determinant of cross-reactive potency. For both Victoria- and Yamagata-like viruses carrying positively charged residues at position 131, antibodies matched to the other positively charged conformation consistently ranked second in potency. In both cases, NQ-purified antibodies, carrying the uncharged ancestral asparagine, provided the weakest neutralisation of viruses with positively charged residues at position 131. This pattern was independently supported by the mutagenesis data (Figure 5). On a B/Yamagata backbone carrying lysine at position 131, K131H (positive to positive) was the least disruptive substitution, while K131N (positive to uncharged) produced a larger reduction in neutralisation. Together, these data demonstrate that antibodies targeting one positively charged residue at position 131 (H or K) cross-neutralise viruses carrying the other positively charged residue more effectively than viruses carrying the uncharged ancestral asparagine.

This transition from an uncharged to a positively charged residue had two consequences. First, it would have enabled cross-recognition by the dominant HH-directed antibody population, since antibodies targeting H at position 131 cross-neutralise lysine carrying viruses via shared positive charge. Second, the timing of this transition, occurring within five years of B/Yamagata’s disappearance, is consistent with a model in which B/Yamagata’s own antigenic evolution at this epitope increased its susceptibility to cross-neutralisation by Victoria-raised immunity. While the population-level asymmetry documented in Figures 1 and 2 reflects the combined effects of multiple epitopes across the HA, the 120-loop data identify a specific molecular mechanism, namely charge-dependent cross-recognition at position 131, that would have contributed to the increasing cross-lineage immune pressure on B/Yamagata in the years preceding its extinction.

## Discussion

This study provides population-level evidence that asymmetric cross-lineage immunity preceded and likely contributed to the disappearance of the influenza B/Yamagata lineage in 2020. Across three independent blood donor cohorts, spanning the period before and after B/Yamagata’s extinction, we found that population immunity was disproportionately focused on B/Yamagata viruses, and that this asymmetry persisted, and initially widened, even after B/Yamagata ceased circulating, consistent with antigenic seniority^14,22^. This asymmetry is consistent with differences in lineage evolution, with B/Yamagata maintaining co-circulating clades, while B/Victoria undergoes more antigenic drift^20,23^, as previously modelled by Han and colleagues^13^.

The identification of position 131 within the 120-loop through computational analysis, and its experimental validation through mutagenesis and epitope-specific pulldowns, establishes a molecular framework for understanding cross-lineage antibody recognition in influenza B. Mutagenesis and serological analyses demonstrate that the charge state at position 131 is a key determinant of neutralisation, with the convergent evolution toward positive charge in both lineages providing a mechanistic basis for altered cross-lineage antibody-mediated immune pressure, consistent with observations for other viruses^24–28^.

The temporal evolution of this epitope provides a potential explanation for B/Yamagata’s disappearance. Historically, B/Yamagata carried an uncharged residue at position 131, limiting recognition by antibodies elicited through B/Victoria exposure. However, after 2015, B/Yamagata acquired a positively charged residue at this site, likely as an escape from Yamagata lineage-specific immunity. This change would have increased susceptibility to cross-neutralisation by Victoria-primed antibodies, effectively exposing B/Yamagata to the dominant population immune response. The timing of this antigenic change, shortly before the lineage’s disappearance, supports a model in which evolutionary adaptation at a single epitope contributed to increased immunological vulnerability, acting alongside external factors such as reduced transmission. The interpretation is supported by studies demonstrating that antigenic evolution at discrete residues can reshape population susceptibility over time, with immune-driven mutations altering the balance between escape and cross-reactive recognition^29,30^.

This study has several potential limitations. The cohorts were geographically restricted to Scotland and may not be representative of populations with different exposure histories or vaccination coverage. In addition, increased influenza vaccine uptake during 2020–22^31^ may have contributed to the boosting observed in 2023. Both mechanisms could plausibly contribute, although the absence of distinct vaccinated and unvaccinated groups appears to make this unlikely. Finally, pseudotyped neutralisation assays, although validated for selected strains, may not capture the full breadth of antiviral immunity, particularly neuraminidase-mediated responses^32,33^.

In summary, our findings suggest that the disappearance of B/Yamagata reflects the combined effects of asymmetric population immunity, and the temporal evolution of a shared HA epitope increasing lineage susceptibility to cross-reactive antibodies. These results highlight how interactions between viral evolution and host immunity can reshape pathogen dynamics. The declining B/Yamagata titres within the 2025 cohort however suggest that the antigenic seniority effect that sustained cross-reactive immunity to B/Yamagata is now waning. This has implications for vaccine policy: should a B/Yamagata-like virus re-emerge, the population may be increasingly vulnerable as the memory responses that once maintained cross-protective immunity erode. Ongoing serological monitoring will be needed to determine whether population immunity to B/Yamagata continues to decline and at what point re-emergence risk becomes significant.

## Methods

### Samples

Blood donor samples were obtained from the Scottish National Blood and Transplant Service (SNBTS) collected between 17th of March and 18th of May 2020, January and November 2023, and January and September 2025. Samples were heat-inactivated before serological testing by incubation at 56°C for 30min.

Ethical approval was obtained for the SNBTS anonymous archive - IRAS project numbers 18005 & 283057. SNBTS blood donors gave fully informed consent to virological testing, donation was made under the SNBTS Blood Establishment Authorisation, and the study was approved by the SNBTS Research and Sample Governance Committee.

Ferret sera was provided by the Crick Influenza Centre.

### Pseudotyped influenza virus production

Pseudotyped lentiviruses displaying influenza haemagglutinins (HAs) were produced by transfecting HEK 293T/17 cells (ECCAC, Public Health England,UK) with: 1.0 μg of gag/pol construct, p8.91, 1.5 μg of a luciferase reporter construct (pCSFLW), 500 ng of TMPRSS4-expressing construct, and 1.0 μg of HA glycoprotein-expressing construct. Transfections were performed in 10 mL of DMEM media supplemented with 10% FBS, 1% penicillin-streptomycin, 1% L-Glutamate and incubated for 16 hours at 37°C. To induce virus budding, 1 unit of endogenous neuraminidase (Sigma, USA) was added to 10 mL of fresh media. Culture supernatants were harvested 48 hours post-budding induction, and stored at -20°C.

The influenza strains used for pseudotyped virus production included: B/Lee/1940, B/Victoria/2/1987, B/Yamagata/16/1988, B/Bangkok/163/1990, B/Hong Kong/03/1992, B/Switzerland/4291/1997, B/Hong Kong/330/2001, B/Florida/4/2006, B/Alaska/01/2010, B/Phuket/3073/2013, and B/Washington/02/2019.

The p8.91, pCSFLW, and TMPRSS4-expressing constructs were gifts from Dr. Nigel Temperton. HA-expressing plasmids were either gifts from Dr. Temperton or produced by cloning GeneArt Strings (Thermo Fisher Scientific, USA) into the pI.18 expression vector (also a gift from Dr. Temperton).

### Pseudotyped influenza virus titration

Serial dilutions were made of pseudotyped influenza virus preparations in Corning Costar 96-well plates (Promega, USA). A total of 10^4^ HEK293T/17 cells were added to each well and incubated for 72 hours at 37°C. The cells were then lysed with BrightGlo reagent (Promega, USA). The relative light units (RLU) of the cell lysate were determined using a Glomax luminometer microplate reader (Promega, USA).

### Pseudotype microneutralisation assay (pMN)

Neutralising antibodies were quantified using a pseudotype microneutralisation assay (pMN). Serially diluted sera was added to 96-well Corning Costar plates (Promega, USA) and incubated with 10^6^ RLU pseudotyped influenza virus for 2 hours at 37°C. Each dilution was performed in duplicate, 2 μL of sera was used per replicate.

Following-virus-serum incubation, HEK 293T/17 cells (2.0 x 10^5^ cells/mL) were added to each well and incubated for 72 hours at 37°C. Cells were then lysed with BrightGlo reagent (Promega, USA). The RLU of the cell lysate were determined using a Glomax luminometer microplate reader (Promega, USA).

The 50% inhibitory dilution factor (ID_50_) was defined as the serum concentration that reduced RLU by 50% compared to virus control wells after subtraction of background RLU from cell-only control wells. Two technical replicates were performed for each biological sample to ensure reproducibility.

### Production of B/Beijing/1/1987 and B/Florida/4/2006 influenza virus

Embryonated hens’ eggs (Medeggs Ltd) were incubated at 37.5°C and 50% relative humidity, turning every 45 minutes until day 10 of embryo development. On day 10, eggs were inoculated with 100 μL of virus (diluted in PBS supplemented with 2% Tryptose Phosphate Broth (Merck) and 5% penicillin-streptomycin) into the allantoic cavity and incubated at 34.5°C and 45% relative humidity, upright, no turning for 72 hours. On day 13, eggs were placed at 4°C overnight to kill the embryos. On day 14, approximately 10 mL of allantoic fluid containing the virus was harvested and centrifuged at 1,000 x g for 10 minutes at 4°C, with the supernatant aliquoted and stored at -80°C. Viral titre was assessed by TCID50 in MDCK-SIAT1 cells supplemented with 2 μg/mL N-tosyl-L-phenylalanine chloromethyl ketone (TPCK)-trypsin (Sigma Aldrich).

### Live virus neutralisation assay

MDCK-SIAT1 cells were seeded at 2 × 10^4^ cells per well in 96-well culture-treated plates (Corning) and incubated at 37°C and 5% CO_2_ overnight. The next day, human sera samples were serially diluted using a 2-fold dilution series in duplicate, starting at a 1:10 dilution, in DMEM supplemented with 0.5% FBS and 2 μg/mL TPCK trypsin (Sigma Aldrich). Virus at 400 TCID50/ml in DMEM supplemented with 0.5% FBS and 2 μg/mL TPCK-trypsin (Sigma Aldrich) was added in equal volume to the diluted sera and incubated for 1 hour at 37°C and 5% CO_2_. The confluent MDCK-SIAT1 cells were washed with 100 μL PBS, before the diluted sera and virus were added and left to incubate at 37°C for 72 hours. Media was removed, and the cells were stained with Coomassie Blue stain (2% Coomassie Blue powder (Thermo Scientific), 50% ethanol absolute, 7.5% acetic acid, diluted in water) for 30 minutes to assess for cytopathic effect (CPE). Wells with no visible CPE were considered neutralised. On separate plates, the virus was diluted in a 5-fold dilution series and incubated for 1 hour at 37°C and then added to confluent MDCK-SIAT1 cells for 72 hours and checked for CPE to confirm that accurate titres of infectious virus were added. Infectious viral titre was calculated by the improved Spearman-Kärber method to be between 300-500 TCID50/mL. The neutralisation result for a serum sample was determined to be the mean of the highest dilutions showing complete neutralisation of the virus.

### Antibody Pulldowns

A subset of neutralising antibodies targeting the full-length HA, head domains (AA 69-310), or head domain epitopes of haemagglutinin were purified from donor sera using an ELISA based pull-down method as described in Steventon,R., 2026. Viral peptides were added to 96 well MaxiSorp plates (Thermo-Fisher Scientific) at a concentration of 10 μg/mL and incubated overnight at 4c. Wells were washed with 0.05% PBS-T before subsequently being blocked with PBS/Casein blocking solution (Thermo-Fisher Scientific) for 1 hour at room temperature. 40 μL of donor sera diluted across two wells was incubated for 2 hours at room temperature before removal of non-bound sera was carried out. Elution of bound antibodies was carried out with 3M MgCl_2_. 5 μL of eluted antibodies was used per replicate in a pMN assay as described above.

### Enzyme-linked immunosorbent assay (ELISA)

Antibody responses to epitope variants were measured using ELISAs. Viral epitopes were added to 96 well MaxiSorp plates (Thermo-Fisher Scientific) at a concentration of 10 μg/mL and incubated overnight at 4c. Wells were washed with 3x 0.05% PBS-T before subsequently being blocked with PBS/Casein blocking solution (Thermo-Fisher Scientific) for 1 hour at room temperature. Donor sera diluted in PBS/casein 1:100 were added to the plate in triplicate. Plates were incubated for 2 hours at room temperature before being washed with 3x 0.05% PBS-T before secondary antibody goat anti-human IgG y chain specific conjugated to alkaline phosphatase (Merck,USA) was added at a dilution of 1:5000 in PBS/Casin solution and incubated for 1 hour at room temperature. After a final wash, plates were developed by adding 4-nitrophenyl phosphate substrate in diethanolamine buffer. Optical density was read at 405nm using a a Glomax absorbance microplate reader (Promega, USA). A reference standard comprising of pooled sera on each plate was serially diluted to generate a standard curve. Three blank wells containing PBS/Casein blocking solution only were used as negative controls. The mean OD values of the blank wells was then subtracted from all OD values on each plate before triplicate mean OD were fitted to the standard curve.

### Augmented Bayesian Network and LASSO regression with grouped positions

An Augmented Bayesian Network (ABN) was constructed from the log₁₀-transformed pMN titres of the 2020 cohort (n = 114 sera) against 11 pseudotyped influenza B viruses using the abn package in R. Neutralisation titres for each strain were treated as continuous Gaussian nodes. Network structure was learned using a score-based hill-climbing search with the Bayesian Information Criterion, restricted to a maximum of three parents per node. The resulting directed acyclic graph represents conditional dependencies between strain-specific neutralisation responses, with edge coefficients (β) quantifying the strength of each dependency.

For each sequence position, pairwise amino acid similarity between strains was encoded as symmetric binary matrices. For each virus pair, a value of 1 was assigned if the two strains shared the same amino acid at that position and 0 otherwise. Positions with identical pairwise similarity patterns across all strain pairs were grouped to avoid redundancy, such that each group represented a set of indistinguishable residue-similarity predictors. For each group, the vectors of 36 non-redundant elements were used for analysis.

Neutralisation titres were skewed and included zero or non-neutralising values and therefore did not satisfy assumptions of normality or linearity required for parametric correlation measures. Therefore, similarity between influenza B viruses was quantified separately for the 2020, 2023, and 2025 cohorts as the Spearman rank correlation of neutralisation titres across all sera for each pair of strains. This yielded a symmetric similarity matrix for each cohort, which was converted into a vector of 36 non-redundant pairwise observations for the 9 strains common to all timepoints. This approach characterises the relative similarity of cross-reactive antibody responses between viruses, rather than absolute neutralisation magnitude.

Grouped amino acid similarity vectors were used as predictors of serological similarity in LASSO (L1-penalised) linear regression. The regularisation parameter (λ) was selected using 10-fold cross-validation, applying the 1-SE rule to favour a parsimonious model. To assess generalisability across cohorts, two complementary analyses were performed. In the primary analysis, predictors were selected using a consensus similarity matrix obtained by averaging correlation matrices from two cohorts (e.g. 2020 and 2023) and evaluated in the held-out third cohort (e.g. 2025). In a secondary analysis, predictors were selected from a single cohort and evaluated on the averaged similarity matrix of the remaining two cohorts. For each training dataset, predictors were ranked by absolute LASSO coefficient magnitude, and the top five predictors were carried forward into a second-stage unpenalised linear regression model fitted on the independent hold-out dataset. This separation of selection and estimation provides unbiased effect size estimates. Bootstrap stability selection (1,000 resamples) was performed by resampling the pairwise strain observations with replacement and refitting the LASSO model in each resample. Sequence alignments and correlation matrices were held fixed, and selection frequency was used to rank predictors.

### Evolutionary sequence analysis

Influenza B HA amino acid sequences were downloaded from the GISAID EpiFlu database (accessed September 2025; n = 80,000 sequences, 1940–2025). Sequences were assigned to B/Victoria or B/Yamagata lineages based on phylogenetic clustering. Amino acid frequencies at positions 131, 132, and 137 were calculated per lineage per year to track the evolutionary trajectory of the 120-loop motif.

### Phlyogenetic analysis

A maximum-likelihood phylogenetic tree was inferred from aligned HA nucleotide sequences using IQ-TREE v3 with automatic model selection (best-fit model: K3Pu+F+G4, selected by BIC) and 1,000 ultrafast bootstrap replicates. Terminal nodes were coloured by amino acid identity at HA position 131. Victoria-lineage strain names are shown in blue and Yamagata-lineage strain names in red.

### Statistical analysis

Comparisons of neutralisation titres between cohorts and between viruses were performed using two-tailed Mann-Whitney U tests with Dunn’s correction for multiple comparisons. Spearman rank correlations were used to assess pairwise associations between neutralisation responses. ELISA binding data were compared using the Friedman test. All statistical analyses were performed in Python (v3.12) using scipy (v1.14), scikit-learn (v1.5), and pandas (v2.2). A significance threshold of p < 0·05 was used throughout.

## Data Availability

All data produced in the present study are available upon reasonable request to the authors and with the agreement of the Scottish National Blood Transfusion Service (SNBTS).

## References

1. Paget J, Caini S, Del Riccio M, van Waarden W, Meijer A. Has influenza B/Yamagata become extinct and what implications might this have for quadrivalent influenza vaccines? Euro Surveill 2022;27:2200753.

2. Vajo Z, Torzsa P. Extinction of the Influenza B Yamagata Line during the COVID Pandemic – Implications for Vaccine Composition. Viruses 2022;14:1745.

3. Caini S, Huang QS, Ciblak MA, et al. Epidemiological and virological characteristics of influenza B: results of the Global Influenza B Study. Influenza Other Respir Viruses 2015;9(Suppl 1):3–12.

4. Caini S, Kusznierz G, Garate VV, et al. The epidemiological signature of influenza B virus and its B/Victoria and B/Yamagata lineages in the 21st century. PLoS One 2019;14:e0222381.

5. Caini S, Meijer A, Nunes MC, et al. Probable extinction of influenza B/Yamagata and its public health implications: a systematic literature review and assessment of global surveillance databases. Lancet Microbe 2024;5:100851.

6. Fisman D, Pérez-Rubio A, Postma M, Smith DS, Mould-Quevedo J. Maintaining the value of influenza vaccination - the shift from quadrivalent to trivalent vaccines: an expert review. Expert Rev Vaccines 2025;24:499–508.

7. Rosu ME, Lexmond P, Bestebroer TM, et al. Substitutions near the HA receptor binding site explain the origin and major antigenic change of the B/Victoria and B/Yamagata lineages. Proc Natl Acad Sci USA 2022;119:e2211616119.

8. Shaw MW, Xu X, Li Y, et al. Reappearance and global spread of variants of influenza B/Victoria/2/87 lineage viruses in the 2000-2001 and 2001-2002 seasons. Virology 2002;303:1–8.

9. Ducatez MF, Webster RG, Webby RJ. Animal influenza epidemiology. Vaccine 2008;26(Suppl 4):D67–69.

10. Pekarek MJ, Weaver EA. Existing Evidence for Influenza B Virus Adaptations to Drive Replication in Humans as the Primary Host. Viruses 2023;15:2032.

11. Chen Z, Tsui JL, Gutierrez B, et al. COVID-19 pandemic interventions reshaped the global dispersal of seasonal influenza viruses. Science 2024;386:eadq3003.

12. Marchi S, Bruttini M, Milano G, et al. Prevalence of Influenza B/Yamagata Viruses From Season 2012/2013 to 2021/2022 in Italy as an Indication of a Potential Lineage Extinction. Influenza Other Respir Viruses 2024;18:e13359.

13. Han W, Zeng J, Shi J, et al. Unraveling the mechanism behind the probable extinction of the B/Yamagata lineage of influenza B viruses. Nat Commun 2025;16:10440.

14. Desheva Y, Kudar P, Sergeeva M, et al. The Persistence of Cross-Reactive Immunity to Influenza B/Yamagata Neuraminidase Despite the Disappearance of the Lineage: Structural and Serological Evidence. Int J Mol Sci 2025;26:7476.

15. Liu Y, Tan HX, Koutsakos M, et al. Cross-lineage protection by human antibodies binding the influenza B hemagglutinin. Nat Commun 2019;10:324.

16. Skowronski DM, Hamelin ME, Janjua NZ, et al. Cross-lineage influenza B and heterologous influenza A antibody responses in vaccinated mice: immunologic interactions and B/Yamagata dominance. PLoS One 2012;7:e38929.

17. Carlock MA, Ross TM. A computationally optimized broadly reactive hemagglutinin vaccine elicits neutralizing antibodies against influenza B viruses from both lineages. Sci Rep 2023;13:15911.

18. Page CK, Shepard JD, Ray SD, et al. Neuraminidase-specific antibodies drive differential cross-protection between contemporary FLUBV lineages. Sci Adv 2025;11:eadu3344.

19. Vijaykrishna D, Holmes EC, Joseph U, et al. The contrasting phylodynamics of human influenza B viruses. Elife 2015;4:e05055.

20. Heider A, Wedde M, Durrwald R, Wolff T, Schweiger B. Molecular characterization and evolution dynamics of influenza B viruses circulating in Germany from season 1996/1997 to 2019/2020. Virus Res 2022;322:198926.

21. Steventon R, Stolle L, Gregory R, et al. Development of an ELISA-Based Pulldown Approach for Functional Analysis of Antigen-Specific Antibodies. bioRxiv 2026;706726.

22. Abozeid HH, Gu C, Trifkovic S, Neumann G, Kawaoka Y. Immunity to Influenza B/Yamagata-Lineage Viruses Has Not Waned Since the Disappearance of This Virus Lineage. Influenza Other Respir Viruses 2025;19:e70188.

23. Langat P, Raghwani J, Dudas G, et al. Genome-wide evolutionary dynamics of influenza B viruses on a global scale. PLoS Pathog 2017;13:e1006749.

24. Sinha N, Mohan S, Lipschultz CA, Smith-Gill SJ. Differences in electrostatic properties at antibody-antigen binding sites: implications for specificity and cross-reactivity. Biophys J 2002;83:2946–2968.

25. Plant EP, Manukyan H, Sanchez JL, Laassri M, Ye Z. Immune Pressure on Polymorphous Influenza B Populations Results in Diverse Hemagglutinin Escape Mutants and Lineage Switching. Vaccines 2020;8:125.

26. Thomson EC, Rosen LE, Shepherd JG, et al. Circulating SARS-CoV-2 spike N439K variants maintain fitness while evading antibody-mediated immunity. Cell 2021;184:1171–1187.e20.

27. Marcinkiewicz AL, Brangulis K, Dupuis AP 2nd, et al. Structural evolution of an immune evasion determinant shapes pathogen host tropism. Proc Natl Acad Sci USA 2023;120:e2301549120.

28. Thompson CP, Lourenço J, Walters AA, Obolski U, Edmans M, Palmer DS, Kooblall K, Carnell GW, O’Connor D, Bowden TA, Pybus OG, Pollard AJ, Temperton NJ, Lambe T, Gilbert SC, Gupta S. A naturally protective epitope of limited variability as an influenza vaccine target. Nat Commun 2018;9:3859.

29. Bedford T, Suchard MA, Lemey P, et al. Integrating influenza antigenic dynamics with molecular evolution. Elife 2014;3:e01914.

30. Kirkpatrick E, Henry C, McMahon M, et al. Characterization of Novel Cross-Reactive Influenza B Virus Hemagglutinin Head Specific Antibodies That Lack Hemagglutination Inhibition Activity. J Virol 2020;94:e01185–20.

31. Kong G, Lim NA, Chin YH, Ng YPM, Amin Z. Effect of COVID-19 Pandemic on Influenza Vaccination Intention: A Meta-Analysis and Systematic Review. Vaccines 2022;10:606.

32. Dutta A, Huang CT, Lin CY, et al. Sterilizing immunity to influenza virus infection requires local antigen-specific T cell response in the lungs. Sci Rep 2016;6:32973.

33. Zhang X, Ross TM. Anti-neuraminidase immunity in the combat against influenza. Expert Rev Vaccines 2024;23:474–484.

34. Ni F, Kondrashkina E, Wang Q. Structural basis for the divergent evolution of influenza B virus hemagglutinin. Virology 2013;446:112–122.

