## Supplementary Figures for "Antigenic seniority and convergent haemagglutinin evolution shaped the immune landscape preceding influenza B/Yamagata’s extinction"

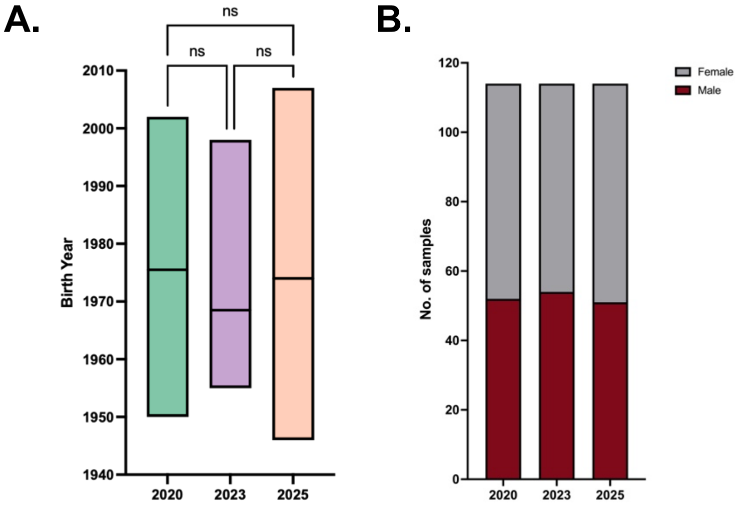


**Supplementary Figure 1. Cohort demographics.** Cohorts did not differ in terms of (A) age or (B) sex. A Kruskal-Wallis test with Dunn’s correction was performed to determine significance.

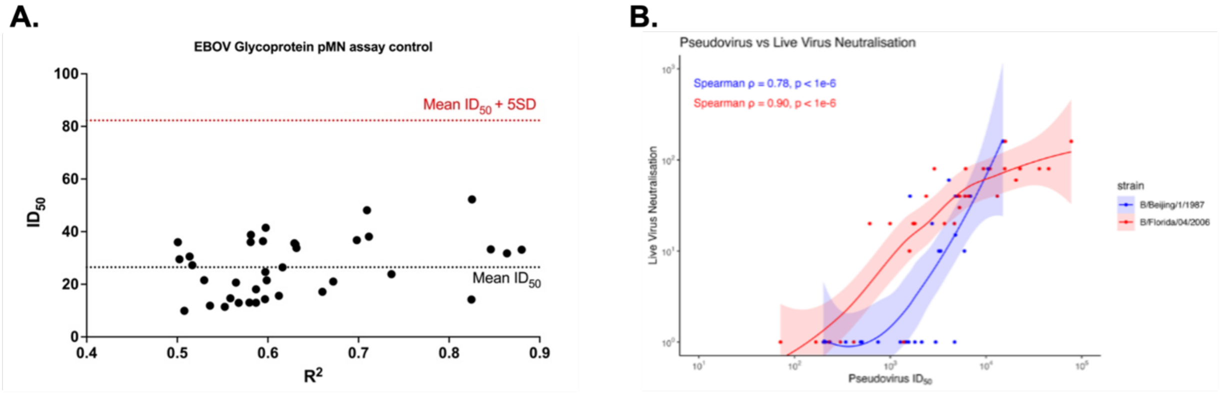


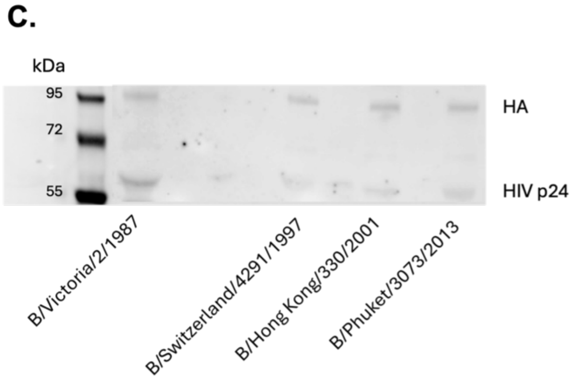


| *Virus* | *Ratio of HA: HIV p24* |
|---|---|
| *B/Victoria/2/1987* | 0.96 |
| *B/Switzerland/4291/1997* | 1.38 |
| *B/Hong Kong/330/2001* | 1.43 |
| *B/Phuket/3073/2013* | 1.12 |

**Supplementary Figure 2: Establishment of pMN assay controls**
(A) Ebola pseudovirus neutralisation assay. 100 sera samples were run against an Ebola pseudovirus, and a seropositivity threshold was determined as five standard deviations above the mean $ID_{50}$. (B) Live Virus Neutralisation correlates strongly with pseudotype neutralisation. 30 sera samples with $ID_{50}$'s against B/Victoria/2/1987 (blue) and B/Florida/4/2006 (red) were tested for neutralisation of live viruses of B/Beijing/1/1987 (99.3% Amino acid similarity to B/Victoria/2/1987) and B/Florida/4/2006. The Spearman rank correlation test showed a significant, strong correlation between pseudotype neutralisation and live virus neutralisation. Negative scores for live virus neutralisation were plotted at $y=1x10^0$ for visualisation purposes. (C) Western blot of HA distribution for selected B/Victoria and B/Yamagata pseudotype viruses, shows no lineage bias in HA distribution. A loading control for the pseudotyped influenza viruswas used.

**A.**

| | B/Lee/1940 | B/Victoria/2/1987 | B/Yamagata/16/1988 | B/Bangkok/163/1990 | B/Hong Kong/03/1992 | B/Switzerland/4291/1997 | B/Hong Kong/330/2001 | B/Florida/4/2006 | B/Alaska/01/2010 | B/Phuket/3073/2013 |
|---|---|---|---|---|---|---|---|---|---|---|
| B/Victoria/2/1987 | <0.0001 | | | | | | | | | |
| B/Yamagata/16/1988 | <0.0001 | >0.9999 | | | | | | | | |
| B/Bangkok/163/1990 | 0.0002 | >0.9999 | 0.0430 | | | | | | | |
| B/Hong Kong/03/1992 | <0.0001 | 0.0268 | >0.9999 | 0.0002 | | | | | | |
| B/Switzerland/4291/1997 | <0.0001 | <0.0001 | 0.0006 | <0.0001 | 0.0961 | | | | | |
| B/Hong Kong/330/2001 | 0.0012 | >0.9999 | 0.0079 | >0.9999 | <0.0001 | <0.0001 | | | | |
| B/Florida/4/2006 | <0.0001 | >0.9999 | >0.9999 | >0.9999 | 0.1253 | <0.0001 | >0.9999 | | | |
| B/Alaska/01/2010 | <0.0001 | >0.9999 | >0.9999 | 0.2561 | >0.9999 | <0.0001 | 0.0589 | >0.9999 | | |
| B/Phuket/3073/2013 | <0.0001 | <0.0001 | 0.0131 | <0.0001 | 0.9258 | >0.9999 | <0.0001 | <0.0001 | 0.0014 | |
| B/Washington/02/2019 | 0.0802 | 0.4498 | <0.0001 | >0.9999 | <0.0001 | <0.0001 | >0.9999 | 0.1132 | 0.0008 | <0.0001 |

**B.**

| | B/Lee/1940 | B/Victoria/2/1987 | B/Yamagata/16/1988 | B/Bangkok/163/1990 | B/Hong Kong/03/1992 | B/Switzerland/4291/1997 | B/Hong Kong/330/2001 | B/Florida/4/2006 | B/Alaska/01/2010 | B/Phuket/3073/2013 |
|---|---|---|---|---|---|---|---|---|---|---|
| B/Victoria/2/1987 | 0.018 | | | | | | | | | |
| B/Yamagata/16/1988 | <0.0001 | <0.0001 | | | | | | | | |
| B/Bangkok/163/1990 | <0.0001 | 0.9723 | 0.0012 | | | | | | | |
| B/Hong Kong/03/1992 | <0.0001 | >0.9999 | 0.0002 | >0.9999 | | | | | | |
| B/Switzerland/4291/1997 | <0.0001 | <0.0001 | >0.9999 | <0.0001 | <0.0001 | | | | | |
| B/Hong Kong/330/2001 | 0.0005 | >0.9999 | <0.0001 | >0.9999 | >0.9999 | <0.0001 | | | | |
| B/Florida/4/2006 | <0.0001 | <0.0001 | >0.9999 | 0.5903 | 0.2051 | 0.011 | 0.0031 | | | |
| B/Alaska/01/2010 | <0.0001 | <0.0001 | <0.0001 | <0.0001 | <0.0001 | 0.0001 | <0.0001 | <0.0001 | | |
| B/Phuket/3073/2013 | <0.0001 | <0.0001 | <0.0001 | <0.0001 | <0.0001 | 0.0028 | <0.0001 | <0.0001 | >0.9999 | |
| B/Washington/02/2019 | 0.0207 | >0.9999 | <0.0001 | 0.9104 | >0.9999 | <0.0001 | >0.9999 | <0.0001 | <0.0001 | <0.0001 |

C.

| | B/Lee/1940 | B/Victoria/2/1987 | B/Yamagata/16/1988 | B/Bangkok/163/1990 | B/Hong Kong/03/1992 | B/Hong Kong/330/2001 | B/Florida/4/2006 | B/Phuket/3073/2013 |
|---|---|---|---|---|---|---|---|---|
| B/Victoria/2/1987 | <0.0001 | | | | | | | |
| B/Yamagata/16/1988 | <0.0001 | >0.9999 | | | | | | |
| B/Bangkok/163/1990 | <0.0001 | >0.9999 | >0.9999 | | | | | |
| B/Hong Kong/03/1992 | <0.0001 | >0.9999 | >0.9999 | >0.9999 | | | | |
| B/Hong Kong/330/2001 | <0.0001 | >0.9999 | >0.9999 | >0.9999 | >0.9999 | | | |
| B/Florida/4/2006 | <0.0001 | >0.9999 | >0.9999 | >0.9999 | >0.9999 | >0.9999 | | |
| B/Phuket/3073/2013 | <0.0001 | <0.0001 | <0.0001 | <0.0001 | <0.0001 | <0.0001 | <0.0001 | |
| B/Washington/02/2019 | <0.0001 | 0.0001 | <0.0001 | <0.0001 | <0.0001 | <0.0001 | <0.0001 | >0.9999 |

**Supplementary Figure 3:** Kruskal-Wallis test with Dunn's correction of **(A)** 2020, **(B)** 2023, and **(C)** 2025 sera. Statistical significance: Statistical significance: *p < 0.05, **p < 0.01, ***p < 0.001, ****p < 0.0001, shown in a green gradient with darker shading representing greater significance.

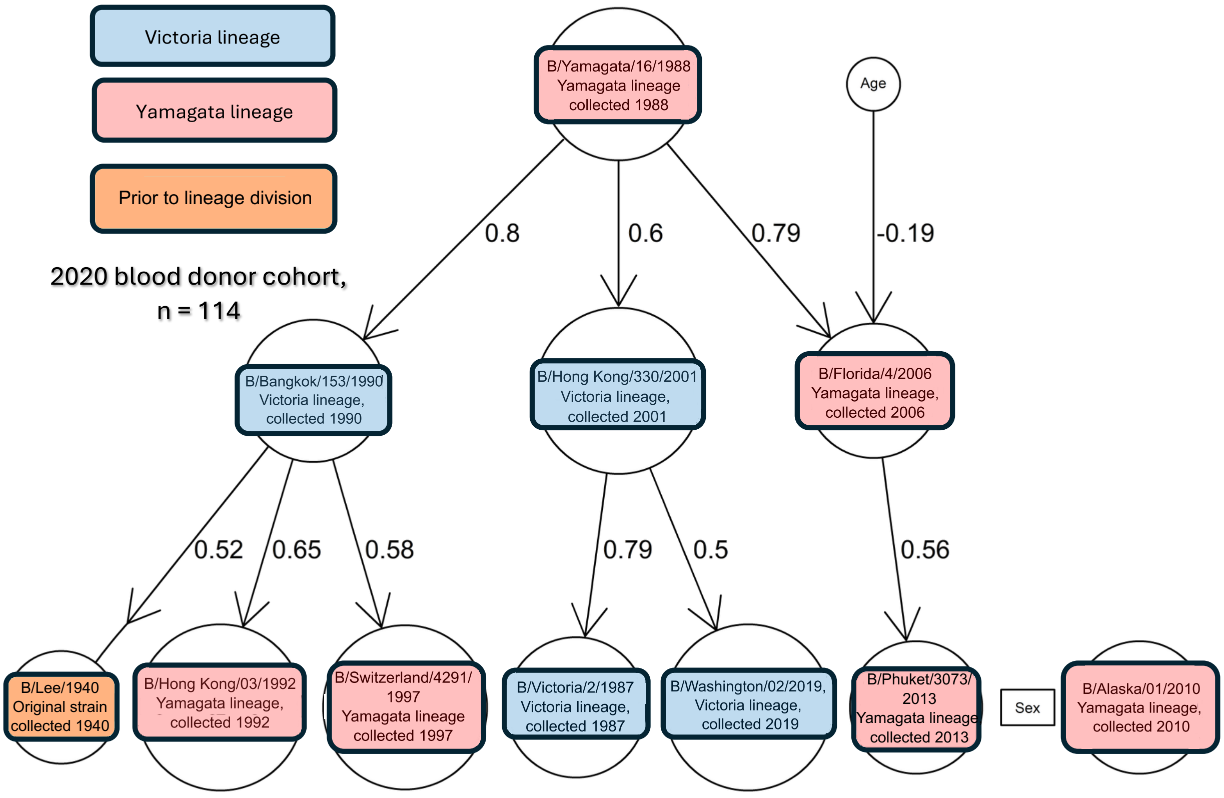


**Supplementary Figure 4. Bayesian network analysis of 2020 neutralisation assay data**
Bayesian network learned from pMN titres of the 2020 donor cohort (n = 114) against 11 influenza B pseudotype viruses. Edge values represent conditional dependency strengths between viruses. Virus lineage is indicated by Victoria (blue), Yamagata (red), and pre-lineage divergence (orange).

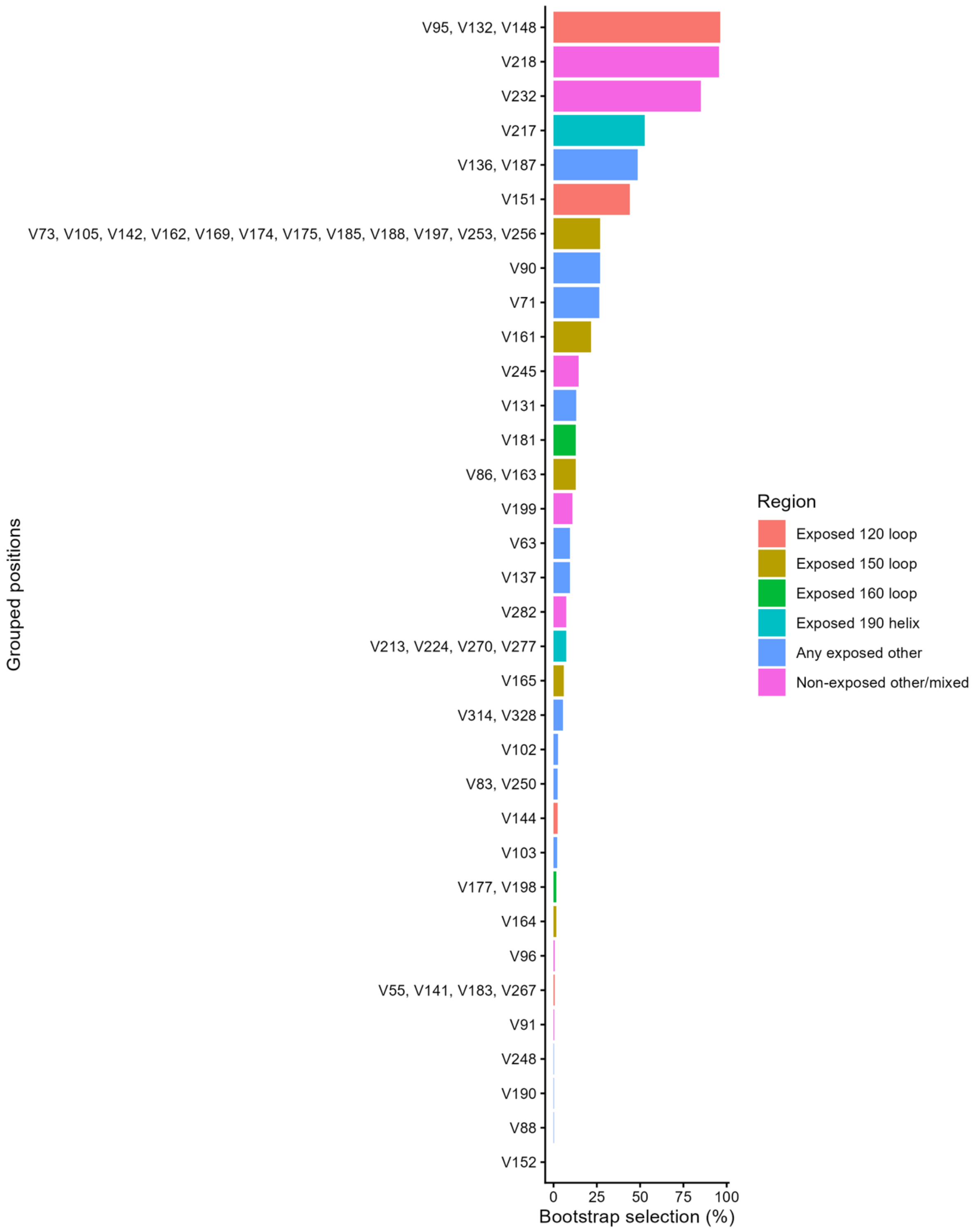


**Supplementary Figure 5. Grouped LASSO bootstrap stability for all 34 independent position groups from the 2020 cohort (11 strains, n = 114 sera).** Perfectly correlated positions were grouped and represented by a single feature. Bars show the percentage of 1,000 bootstrap resamples in which each group was selected, colour-coded by antigenic region based on the first exposed position within the group: 120 loop (salmon), 150 loop (olive), 160 loop (green), 190 helix (cyan), any exposed other (blue), and non-exposed or mixed (pink). Antigenic site boundaries follow Ni et al. (2014): 120 loop (HA1 116-137), 150 loop (HA1 141-150), 160 loop (HA1 162-167), 190 helix (HA1 194-202). Dashed line at 50% for reference.

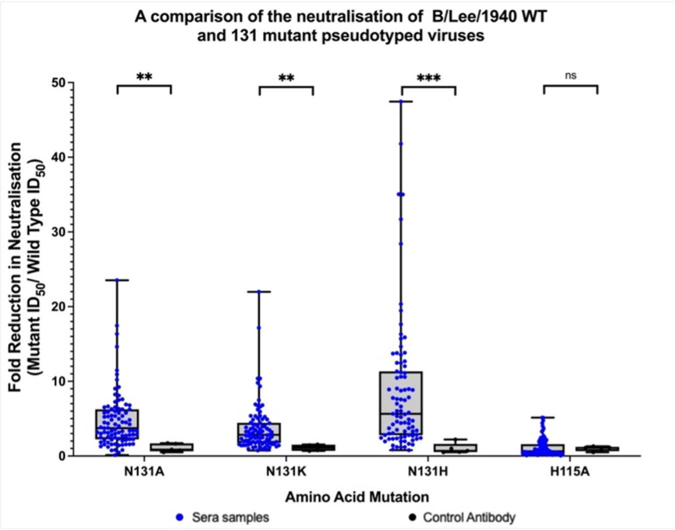


**Supplementary Figure 6: Switching the amino acid present at position 131 results in a significant decrease in the neutralising capacity of the sera samples.** SDM of residue 131 in the B/Lee/1940 background. CR9114 was used as a control monoclonal antibody. 114 sera samples from the 2020 donor cohort were run in pMN assays against mutated pseudotype viruses. Y axis denotes fold reduction in neutralisation in 50% inhibitory dilution factor ($ID_{50}$) units. Statistical analysis was performed using a two-tailed Mann-Whitney U test with Dunn's correction. Asterisks denote statistical significance: $p < 0.05$ (*), $p < 0.01$ (**), $p < 0.001$ (***), $p < 0.0001$ (****).

**Supplementary Tables:**

| Strain | Lineage | Cohorts run against |
|---|---|---|
| B/Lee/1940 | Ancestral | 2020, 2023, 2025 |
| B/Victoria/2/1987 | Victoria | 2020, 2023, 2025 |
| B/Yamagata/16/1988 | Yamagata | 2020, 2023, 2025 |
| B/Bangkok/163/1990 | Victoria | 2020, 2023, 2025 |
| B/Hong Kong/03/1992 | Yamagata | 2020, 2023, 2025 |
| B/Switzerland/4291/1997 | Yamagata | 2020,2023 |
| B/Hong Kong/330/2001 | Victoria | 2020, 2023, 2025 |
| B/Florida/4/2006 | Yamagata | 2020, 2023, 2025 |
| B/Alaska/01/2010 | Yamagata | 2020,2023 |
| B/Phuket/3073/2013 | Yamagata | 2020, 2023, 2025 |
| B/Washington/02/2019 | Victoria | 2020, 2023, 2025 |

**Supplementary Table 1: List of strains used within pMN assays**

| Rank | Position | Amino acid variants | Antigenic region | LASSO | LASSO +Hamming | LASSO +PCoA | Elastic Net | EN +Hamming | EN +PCoA |
|---|---|---|---|---|---|---|---|---|---|
| 1 | 95 | K/R | | 99.5 | 99.5 | 99.2 | 99.7 | 99.5 | 99.3 |
| 2 | 218 | K/N/R/S | 190 helix | 97.9 | 96.7 | 97.7 | 97.9 | 96.7 | 97.9 |
| 3 | 232 | A/V | | 97.1 | 97.2 | 96.2 | 97.1 | 97.2 | 95.6 |
| 4 | 132 | I/V | 120 loop | 96.5 | 96.6 | 95.4 | 99.7 | 99.5 | 99.3 |
| 5 | 136 | N/T | 120 loop | 89.8 | 91 | 91.2 | 90 | 91.1 | 89.5 |
| 6 | 156 | E/G | 150 loop | 88.6 | 90.9 | 89.3 | 88.4 | 90.6 | 88 |
| 7 | 148 | G/R | 120 loop | 79.9 | 85 | 84.6 | 99.7 | 99.5 | 99.2 |
| 8 | 187 | P/S | | 76.7 | 84.7 | 82.3 | 89.7 | 90.9 | 89 |
| 9 | 160 | N/S | 150 loop | 76.4 | 79.1 | 79.3 | 88.4 | 90.6 | 88 |
| 10 | 71 | D/K/N/T | | 55.1 | 38.4 | 54.3 | 54.6 | 37.8 | 51.5 |
| 11 | 151 | E/K/R | 120 loop | 51.9 | 47.5 | 50.4 | 51.4 | 47.1 | 49 |
| 12 | 217 | A/E/K/V | 190 helix | 50.2 | 37.1 | 44.2 | 50.7 | 36.9 | 44.5 |
| 13 | 105 | A/V | | 40.6 | 19.6 | 8.1 | 32.3 | 18.4 | 5 |
| 14 | 162 | A/T | 150 loop | 39.3 | 20.4 | 8.8 | 48.9 | 25.7 | 9.2 |
| 15 | 161 | A/I/V | 150 loop | 36.1 | 28.2 | 34.5 | 36 | 28.2 | 33.6 |
| 16 | 142 | A/T | 120 loop | 34.4 | 16.1 | 5.8 | 39 | 18.5 | 7.3 |
| 17 | 199 | E/G | | 33.2 | 33.9 | 33.8 | 32.7 | 33.5 | 31.2 |
| 18 | 90 | I/K/N/T | | 32.4 | 24.2 | 28.4 | 32.2 | 23.9 | 28 |
| 19 | 169 | A/N | | 31.3 | 15.8 | 7.7 | 48.9 | 25.7 | 9.2 |
| 20 | 73 | F/L | | 28.5 | 9.6 | 4.7 | 39.3 | 21.4 | 6.1 |

**Supplementary Table 2**: Model comparison showing bootstrap stability across 6 model variants (LASSO, LASSO+Hamming, LASSO+PCoA, Elastic Net, EN+Hamming, EN+PCoA) for selected positions.

| Rank (full HA) | Position | Amino acid variants | Domain | Spearman ρ | p-value |
|---|---|---|---|---|---|
| Top HA1 positions for comparison | | | | | |
| 1 | 148 | G/R | HA1 | 0.5765 | 0.000004 |
| 3 | 95 | A/K/R | HA1 | 0.4772 | 0.000229 |
| 4 | 132 | I/N/V | HA1 | 0.4772 | 0.000229 |
| 5 | 63 | E/K/R/T | HA1 | 0.4531 | 0.000513 |
| 6 | 90 | G/I/K/N/T | HA1 | 0.427 | 0.001149 |
| All HA2 positions | | | | | |
| 2 | 520 | D/N | HA2 | 0.5765 | 0.000004 |
| 13 | 570 | I/V | HA2 | 0.2858 | 0.034416 |
| 30 | 566 | I/L | HA2 | 0.1629 | 0.234617 |
| 31 | 494 | D/E | HA2 | 0.1565 | 0.253783 |
| 33 | 522 | G/R | HA2 | 0.1412 | 0.303979 |
| 34 | 574 | I/V | HA2 | 0.1412 | 0.303979 |
| 185 | 416 | S/Y | HA2 | 0.0683 | 0.620020 |
| 186 | 433 | D/N | HA2 | 0.0683 | 0.620020 |
| 187 | 437 | D/N | HA2 | 0.0683 | 0.620020 |
| 188 | 523 | D/E | HA2) | 0.0683 | 0.620020 |

Supplementary Table 3. Full-length HA analysis showing the top HA1 positions for comparison alongside all 10 variable HA2 positions. Only 2 of 10 HA2 positions reach significance.
